# Robust Mixed Model Association Test for Gene-Environment Interactions

**DOI:** 10.1101/2025.10.01.25336808

**Authors:** Mengyu Zhang, Jingxian Tang, Michael R. Brown, Alanna C. Morrison, Eric Boerwinkle, Alisa K. Manning, Ching-Ti Liu, Han Chen

**Affiliations:** Department of Biostatistics and Data Science, The University of Texas Health Science Center at Houston; Department of Biostatistics, Boston University School of Public Health; Human Genetics Center, Department of Epidemiology, The University of Texas Health Science Center at Houston; Human Genetics Center, Department of Epidemiology, The University of Texas Health Science Center at Houston, Human Genome Sequencing Center, Baylor College of Medicine; Programs in Metabolism and Medical & Population Genetics, Broad Institute of MIT and Harvard, Department of Medicine, Harvard Medical School, Clinical and Translational Epidemiology Unit, Massachusetts General Hospital

**Keywords:** Large-scale, gene-environment interaction, robust association test, Huber-White sandwich estimator, linear mixed model

## Abstract

Linear mixed models (LMMs) are widely used in gene-environment interaction (GEI) studies to account for population structure and relatedness. However, genome-wide GEI tests using LMMs are computationally intensive, and model-based tests can yield inflated type I error rates when environmental main effects are misspecified. While robust inference methods exist for unrelated samples, challenges remain for related individuals. A common workaround is a two-step approach that first adjusts for relatedness via an LMM and then uses residuals in a standard linear model, but its validity for GEI studies is unclear. We propose a robust mixed model association test (RoM) for large-scale GEI analysis in related samples. RoM uses the Huber-White sandwich estimator and offers efficient computation, scaling linearly with sample size when cluster sizes are bounded. Simulations show that RoM achieves better type I error control at genome-wide significance levels than both the two-step method and alternative strategies. We apply RoM to GEI analyses of waist-hip ratio (WHR) with BMI using data from the Framingham Heart Study (7,264 related individuals), ARIC (9,312 individuals with repeated measures), and WHR with sex using data from UK Biobank (407,068 related individuals), confirming robust error control and comparable signal detection.

## 1 Introduction

Many complex diseases stem from interaction between gene and non-genetic risk factors including lifestyle, environmental exposure, physical condition, etc., and how each factor contributes to the disease can vary from person to person. For example, genetic variants in certain genes such as *TCF7L2* (Cauchi & Froguel 2008), *KCNJ11* (Haghvirdizadeh et al. 2015), and *PPARG* (Janani & Kumari 2015) are known to regulates insulin production, secretion and sensitivity and so affect the risk of developing type 2 diabetes. However, lifestyle factors such as sedentary lifestyles and unhealthy diets can also contribute to the development of type 2 diabetes (Ali 2013). Similarly, a higher incidence of developing Parkinson’s disease has been linked to genetic variants in genes such as *LRRK2, SNCA*, and *GBA* and environmental exposures such as exposure to pesticides, herbicides, and certain metals (Goldman 2014). Gene-environment interaction (GEI) tests focus on genetic alleles that modify the effect of environmental exposures on complex diseases and traits, which help us understand their full genetic architecture. In addition, joint tests of genetic main effects and GEI aim to identify genetic associations after accounting for possible heterogeneous effects in different environmental exposure strata. They have been successfully applied to large-scale genome-wide association studies (GWAS) on fasting glycemic traits (Manning et al. 2012), blood lipids traits (Kilpeläinen et al. 2019), and blood pressure (Nagarajan et al. 2025). GEI models and statistical analysis provide crucial insights into etiology of complex disease (McAllister et al. 2017) by identifying exposures that have different effect on disease for people with different genotypes or genotype that have different impact on disease among people in different environmental exposures.

Linear mixed model (LMM) is a powerful tool for genetic association tests that take into account population structure and relatedness (Zhou & Stephens 2012, Loh et al. 2015, Mbatchou et al. 2021). However, conducting genome-wide single-variant genetic and GEI tests using standard test statistics with LMM for large sample sizes can be computationally burdensome (Bi & Lee 2021). First, large matrix computation can be memory-intensive and slow down computation. Second, classical models require separate model fitting for each SNP being tested, regardless of whether a Wald test or a score test is used. To address these issues, MAGEE (Wang et al. 2020) was developed to improve efficiency by fitting only one global null model and conducting GEI tests after projecting out genetic main effects. However, its model-based hypothesis test makes it susceptible to model misspecification.

Model misspecification is a common problem in statistical modeling that would lead to biased coefficient estimate and inconsistent standard error (Tchetgen & Kraft 2011). It can arise from the exclusion of important variables, such as sex and age, or from an incorrect functional form. In genetic association studies, misspecification of the model refers to the situation in which the model inadequately captures the true relationship between genotype and phenotype. Furthermore, pooling different populations without appropriate adjustments can also lead to model misspecification in genetic association studies (Pham et al. 2023). A specific example of an incorrect functional form is the mis-specification of nonlinear environmental main effects as linear effects which can result in inflated type I error rates in gene by environment association tests. Cornelis et al. (2012) implemented binary BMI to handle the type I error inflation in the gene by the BMI interaction association test, resulting from misspecifying the main effect of continuous BMI. Voorman et al. (2011) found that even a slight model misspecification could result in a significant inflation of model-based statistics, but they did not observe such an inflation for robust inference using Huber-White standard error.

Sandwich estimator is usually used in Generalized Estimating Equation (GEE), where the correlation structure is handled by a working correlation structure, enabling the generation of robust standard errors even when this structure is incorrectly specified. GEE are robust for handling clustered data, yet they fall short in modeling intricate kinship correlations in genetic study. Additionally, conducting genome-wide interaction tests with GEE is computationally intensive because each genetic variant requires fitting an individual model. Therefore, Robust inference procedures have been widely applied in GEI studies for unrelated samples (Almli et al. 2014, Kerin & Marchini 2020). Westerman et al. (2021) also developed the software GEM using a robust approach under a linear model or logistic linear model in millions of unrelated samples, and it performed well on type I error control. However, the challenge remains for related samples. A two-step procedure, in which a linear mixed model is first fitted to account for sample relatedness and the resulting residuals are used as outcomes in a subsequent linear model, has been proposed (Aulchenko et al. 2007, Sofer et al. 2019) and widely applied in GEI study Aulchenko et al. (2010). While this approach provides a practical solution for decorrelating samples, its performance for heritable environmental exposures (e.g., BMI) and in the presence of potentially misspecified environmental main effects in GEI studies has not yet been systematically evaluated. Finally, Freedman has proposed a sandwich estimator for related samples with variances “adjusted for clustering” (Freedman 2006), but it has not yet been applied to GEI studies.

We propose a robust mixed model association test (RoM) for large-scale GEI in related samples and repeated measurements. We implemented this single variant analysis based on the Huber-White sandwich estimator and it has an efficient implementation that scales linearly with the sample size when cluster sizes are bounded. The paper is organized as follows. In Section 2, we introduce our models, test statistic, and four alternative methods. We also present a comprehensive simulation study to evaluate Type I error rate control of five strategies and real data analyses in three studies. Specifically, we examine the gene-body mass index (BMI) on waist-hip ratio (WHR) in 7,264 related individuals spanning three generations in the Framingham Heart Study (FHS). We further investigate the same interaction effect on WHR among 9,312 independent European Americans in the Atherosclerosis Risk in Communities (ARIC) study, each with up to six repeated measurements. Lastly, we evaluate the gene-sex interaction on WHR among both related (up to 407,068) and unrelated (up to 275,478) White British participants in the UK Biobank. Section 3 details the results from the simulations and real data analysis, and Section 4 discusses our findings and implications.

## 2 Materials and Method

### 2.1 Linear mixed models (LMMs)

For single-variant test, RoM considers the following linear mixed model for related individuals

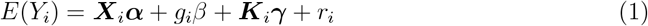

where *Y*_*i*_ is the phenotype for subject *i, i* = 1, …, *N*. ***X***_*i*_ is a 1 × *p* vector of covariates for subject *i, g*_*i*_ is the genotype of a variant for subject *i*, ***K***_*i*_ is a 1 × *q* vector of GEI terms for subject *i* and environmental exposures of interest are a subset of covariates. ***α*** is covariate effects, *β* is genetic main effect, ***γ*** is GEI effect, and *r*_*i*_ is random effect of subject 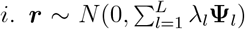 is a vector with elements *r*_*i*_, where *λ*_*l*_ is the variance component parameter for *L* random effects and **Ψ**_*l*_ are *N* × *N* relatedness matrices. Under the null of no GEI effect *H*_0_: ***γ*** = 0, the model becomes

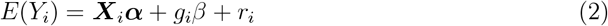

Under the null hypothesis of no genetic main effect or GEI effect for the joint test *H*_0_: *β* = 0, ***γ*** = **0**, the model becomes

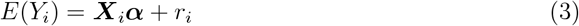

### 2.2 Robust association tests

With misspecified nonlinear environmental effect, the model-based estimator may underestimate the variance of interaction effect, leading to biased standard errors and inflated Type I error rates. In the case of family data, where environmental exposures is heritable like BMI, the problem of inflated Type I error rates due to model misspecification can be more severe. Similar issue can occur when analyze longitudinal/cluster data if the correlation structure is incorrectly specified. Sandwich estimators can be used to address this issue in most cases by sandwiching the estimator of the covariance matrix of samples between two estimators of the variance of the coefficients. Sandwich estimators are robust because this method does not rely on assumptions about the distribution of the errors or the functional form of the regression model. Therefore, robust standard errors are insensitive to misspecification of the correlation structure within individual or clustered measurements. In the description of the model, the matrix containing information of sample covariance matrix is referred to as the”meat” matrix ***M***, and the estimated variance of coefficients is referred to as the “bread” matrix ***B***. Inspired by the idea of sandwich estimator, we propose an efficient single-variant robust score test for GEI effect and joint effect of variant and GEI.

As previously mentioned, conducting single-variant tests using classical models can be computationally intensive due to the requirement of fitting separate models for each SNP being tested; therefore, to alleviate this burden, we first fit a null mixed model with GMMAT (Chen et al. 2016) under the null hypothesis of no interaction or joint effect as Equation (3)

Then performing robust score tests by constructing the “bread” matrix 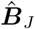 and 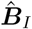 for joint and interaction test as

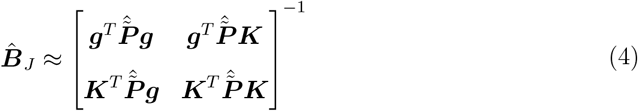

and

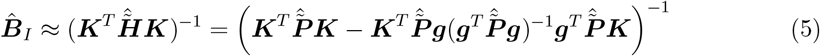

respectively, where ***g*** = (*g*_1_, …, *g*_*n*_)^*T*^ is a *n* × 1 vector representing genotypes for a certain variant of all subjects and ***K*** is a *n* × *q* matrix representing GEI terms for a certain variant of all subjects. 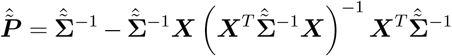 and 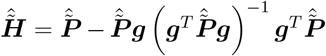 are estimated projection matrices. 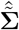 is the covariance matrix for samples estimated from null model in Equation (3). The “meat” matrix for the joint test is

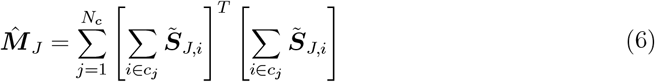

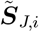 is the score vector for subject *i. c*_*j*_ is set of individuals belonging to clusters *j. N*_*c*_ is the total number of clusters. The “meat” matrix for GEI test 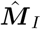 is the corresponding block of 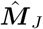. Details can be found in Supplementary Note A.

Therefore, the Huber-White sandwich estimator of variance for joint test is

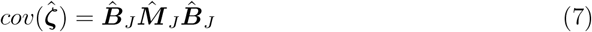

where 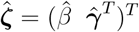, and robust variance for interaction test is

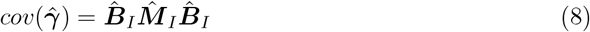

Estimate of genetic main effect and interaction effect is computed by

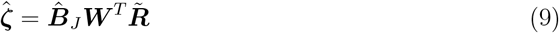

where ***W*** = (***g, K***), and 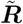 is the scaled residuals from null model in Equation (3). The Score Test statistic for joint test is

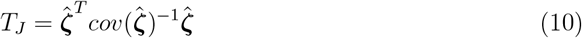

which follows 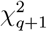 under the null *H*_0_: ***ζ*** = **0**, and the Score Test statistic for interaction test is

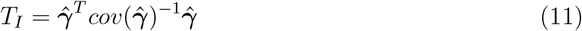

which follows 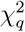 under the null *H*_0_: ***γ*** = 0.

RoM is computationally efficient on association testing. It avoids fitting models repeatedly for each SNP being tested. The computation of scores and residuals involves projection of projection matrix, and the complexity of multiplication of two *N* × *N* matrices is *O*(*N* ^3^) which will slow down the algorithm dramatically. To address this issue, we compute the residuals without calculating projection matrix explicitly. Additionally, the largest cluster or family size is bounded compared to the total sample size, and the “meat” matrix can be easily computed with the membership matrix 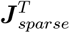. Eventually, the computation complexity of RoM is *O*(*N*). More details can be found in Supplementary Note B.

### 2.3 Simulation Studies

For the purpose of exploring how model misspecification lead to inflated type I error in gene-environment interaction tests for common variants in family studies, we simulated independent 1,000,000 variants (MAF *>* 0.05) and 14,750 families with 85,000 individuals in total. There are 12,750 nuclear families with 2 parents and 2 offspring, and 2,000 extended families with 3 offspring for each generation as shown in Supplementary Figure S1. Genotypes of 35,500 founders were generated from Binomial distribution, and each offspring randomly inherited one haplotype from each parent. Kinship matrix was computed using R package kinship2 (Sinnwell et al. 2014). The simulation was conducted on the Lonestar6 system at the Texas Advanced Computing Center (TACC) The University of Texas at Austin.

We simulated 2,000 continuous traits replicates from

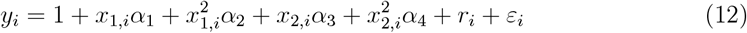

where *x*_1_ *∼ N* (0, 2**Ψ**) and *x*_2_ *∼ N* (0, 2) are environment terms, **Ψ** is kinship matrix, ***r*** *∼ N* (0, 2**Ψ**), and random error *ε*_*i*_ *∼ N* (0, 1). Effect size *α*_*j*_, *j* = 1, 2, 3, 4 were decided by fixing the variance proportion of traits for environment terms to 0.1, 0.05, 0.1, 0.05, respectively. Therefore, the variance explained by 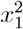 is 0.05. A misspecified model with the dropped non-linear environment effect is

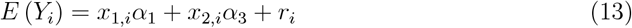

We aimed to investigate two scenarios: one in which the environmental exposure is heritable (*x*_1_), and another in which it is non-heritable (*x*_2_). The performance of the RoM was assessed in both scenarios and compared against alternative strategies, as outlined below.

#### 2.3.1 LMM-model-based strategy (LMM-MB)

LMM-model-based strategy serves as a negative control that is expected to show inflated type I error in the presence of environment main effect misspecification. LMM-MB builds a linear mixed null model as in Equation (3) and conducts the GEI and joint tests as in Equation (14) and Equation (15), respectively.

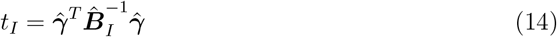

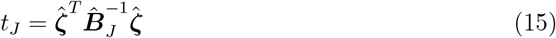

#### 2.3.2 LMM-individual-cluster-based strategy (LMM-ICB)

The individual-cluster-based (ICB) sandwich estimator is a straightforward extension of the sandwich estimator typically used for unrelated individuals. However, when applied to family studies or related individuals, the clustering is inherently misspecified, as each individual is treated as an independent cluster. To evaluate its empirical performance, we consider the LMM-individual-cluster-based strategy. This strategy involves fitting the same linear mixed null model and testing using individual-cluster-based robust variance estimator. As a result, the “meat” matrix changes from Equation (6) to

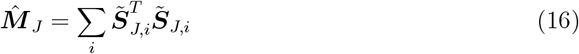

#### 2.3.3 Two-step-individual-cluster-based strategy (Two-step-ICB)

As described in the introduction, a two-step approach has been applied in practice. It de-correlates the study samples by fitting a linear mixed model to account for sample relatedness and treating the residuals as the outcome in a linear model in the second step. To assess its empirical performance, we set up the two-step-individual-cluster-based strategy that fits model with two-step approach and conducts association test using individual-cluster-based robust variance estimator.

#### 2.3.4 LM-individual-cluster-based strategy (LM-ICB)

LM-individual-cluster-based strategy is a negative control to two-step-ICB strategy which fits a linear null model on covariates and use individual clusters when computing “meat” matrix.

### 2.4 Gene-BMI interaction studies of waist-hip ratio in FHS and ARIC

We conducted gene-BMI interaction study using data from FHS and ARIC to show the performance of different test strategies mentioned above on controlling inflated type I error in cluster samples when model is misspecified.

FHS (Mahmood et al. 2014) began in 1948 with the recruitment of an original cohort of 5,209 men and women (mean age 44 years; 55 percent women). In 1971 a second generation of study participants was enrolled; this cohort (mean age 37 years; 52% women) consisted of 5,124 children and spouses of children of the original cohort. A third-generation cohort of 4,095 children of offspring cohort participants (mean age 40 years; 53 percent women) was enrolled in 2002-2005 and are seen every 4 to 8 years. At each clinic visit, a medical history was obtained with a focus on cardiovascular content, and participants underwent a physical examination including measurement of height and weight from which BMI was calculated. WHR was taken from the fourth cohort examination of the offspring cohorts (refer to appointment time). 7,264 individuals and 1,366 families in FHS were included in the real data analysis. Genotype imputation was performed using SHARe 550K SNP array data (Affymetrix 500K mapping array plus Affymetrix 50K supplemental array) as input, conducted in May 2020 through the TOPMed Imputation Server (Fuchsberger et al. 2014, Das et al. 2016, Taliun et al. 2021) (publicly released April 2020). Variant positions are reported based on genome build GRCh38/hg38. The dbGaP study accession number is phs000342.v16.p10.

The ARIC study is a population-based prospective cohort study of cardiovascular disease sponsored by the National Heart, Lung, and Blood Institute (NHLBI). ARIC included 15,792 individuals, predominantly European American and African American, aged 45-64 years at baseline (1987-89), chosen by probability sampling from four US communities. Cohort members completed three additional triennial follow-up examinations, a fifth exam in 2011-2013, a sixth exam in 2016-2017 and several subsequent exams. BMI and WHR were collected for each visit. The ARIC study has been described in detail previously (Wright et al. 2021). 9,312 European Americans and about 38,000 observations measured spanning up to 6 visits were included in our analysis. ARIC samples were genotyped on the Affymetrix 6.0 genotyping array and imputed to the TOPMed R2 reference panel using the TOPMed Imputation Server (Fuchsberger et al. 2014, Das et al. 2016, Taliun et al. 2021) powered by BioData Catalyst. Prior to imputation, genotyped samples were excluded if they failed to meet the following criteria: minor allele frequency *>* 0.01, sample missingness *<* 0.05, and Hardy-Weinberg Equilibrium *>* 0.00001.

Gene-BMI interaction on the quantitative trait waist-to-hip ratio (WHR) was tested on 22 chromosomes with about 9 million common variants (MAF *>* 0.01) from imputed genotypes with adjustments including age, biological sex, age^2^, sex×age, sex×age^2^, and 10 genetics principal components (PCs). Additionally, two indicator variables of field center, centerw (whether in Washington County, Maryland) and centerf (whether in Forsyth County, North Carolina), were adjusted in ARIC. The indicator variables, such as offspring and gen3 for generations ID, were adjusted in FHS.

We investigated the performance of six models on the real data. We considered model-based statistics in LMM (BMI-LMM-MB), in LMM with quadratic BMI term (BMI2-LMM-MB) and in LMM with random slope of BMI (randBMI-LMM-MB). In addition to model-based statistics, robust family-cluster-based statistics in LMM (BMI-RoM), robust family-cluster-based statistics in LMM with random slope of BMI(randBMI-RoM) and robust individual-clustered-based statistics in two-step approach with random slope of BMI (randBMI-two-step-ICB) were included.

### 2.5 Gene-sex interaction studies of waist-hip ratio in UK Biobank

To show the capability of RoM on identifying the signals for large sample size study, we conducted a gene-sex interaction analysis using UK Biobank participants of White British ancestry. To illustrate the power gained and lower signal p values by including related individuals, we applied RoM to two datasets: one with 275,478 unrelated White British participants and another involving related White British participants (in total N = 407,068) in comparison to the analysis of unrelated individuals in Westerman et al. (2021). Array-typed genotypes were transformed to GDS format using R package SeqArray with filter minor allele frequency (MAF) *>* 0.001. Following the methodology in Westerman et al. (2021), WHR was transformed using an inverse-normal approach prior to the analysis, and the model included additional covariates such as age, age^2^, the top 10 genetic principal components, BMI, and a gene-BMI interaction. GEI test and joint test results were annotated using the FUMA web tool (Watanabe et al. 2017). We defined independent significant SNPs as those reached P-value = 5 × 10^−8^ and are independent at *r*^2^ *<* 0.6. Lead SNP was defined as those with a P-value of ≤ 5 × 10^−8^ and selected from independent significant SNPs which are independent each other at *r*_2_ *<* 0.1. Independent SNPs were merged into a single genomic locus using a 250kb window.

## 3 Results

### 3.1 Type I error control simulation

We evaluated inflation of type I error rates from three perspectives: genomic inflation factor (GIF) *λ*_*gc*_, QQ plots, and empirical type I error rates. GIF is the ratio of median of observed statistics and median of test statistics under the null. It helps detect whether the association signals in a genetic study are inflated due to population structure or systematic bias in the test statistics of each SNP.

The violin and QQ plots in Figure 1 show consistent results of GEI test and joint test. Type I error rate for model-based statistics (LMM-MB) is inflated when nonlinear environment effect is misspecified in the model. RoM has robust performance on type I error rate control. Two-step approach that is of the most interest along with other individual-cluster-based statistics did not perform well on producing well-calibrated p values, except for the GEI test of not heritable environment factor (*x*_2_) in Figure 1g. Gene by heritable environment factor (*x*_1_) produced more deflated joint test results (*x*_1_ Joint *λ*_*GC*_ = 0.767 in Figure 1f; *x*_2_ Joint *λ*_*GC*_ = 0.817 in Figure 1h). Empirical type I error rates summarized in Table 1 give the numerical results depicted in the plots.

**Figure 1:**
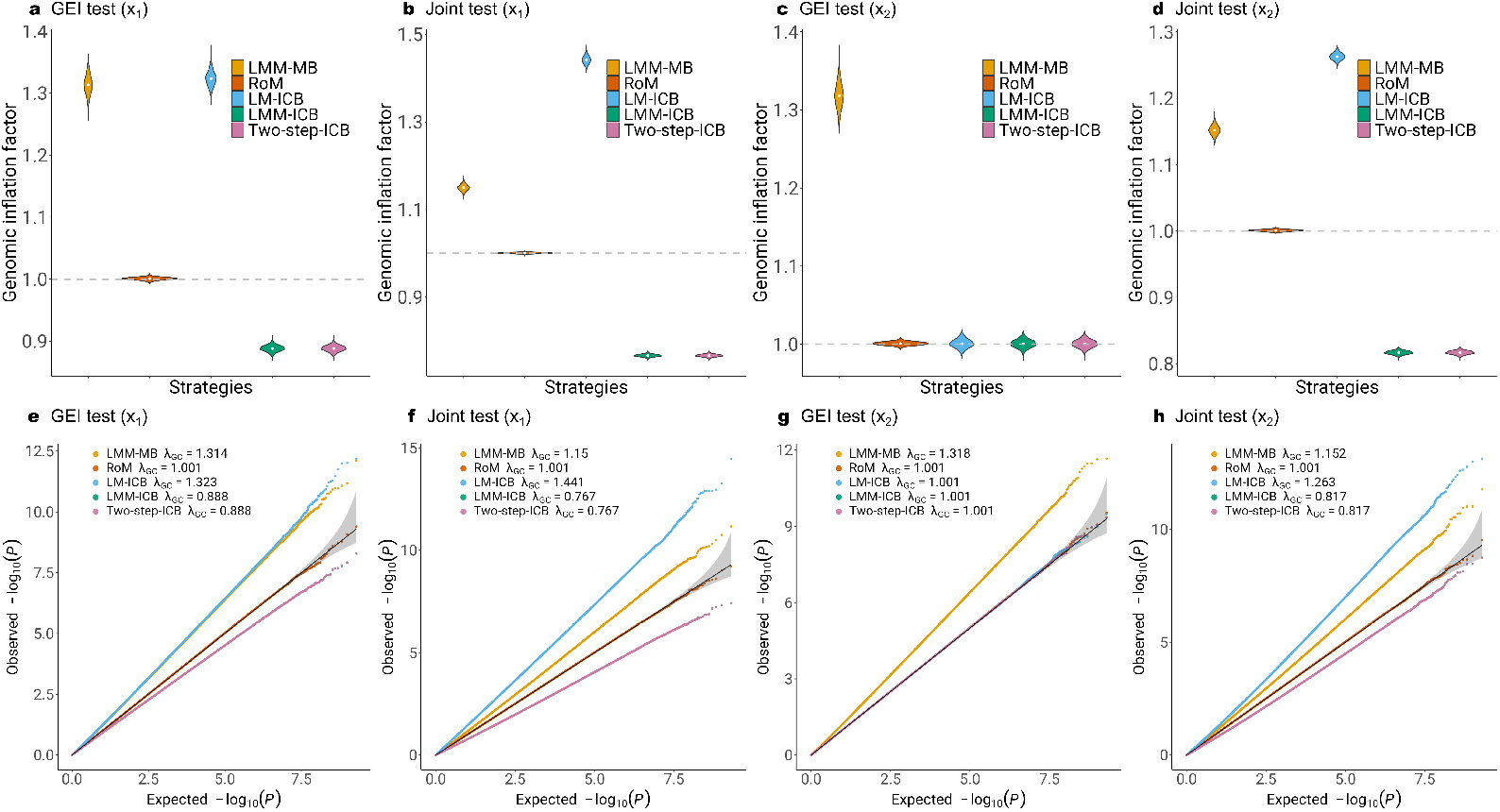
Comparison of GIF and QQ plots of RoM and four alternative strategies under the null hypothesis of no genetic effect or GEI effect for a sample size of 85,000. a-d) Genomic inflation factors over 2,000 replicates. e-h) QQ plots of 2 billion SNPS from 2,000 replicates combined. LMM stands for linear mixed model. LM stands for linear model. Two-step stands for two-step approach applying LMM in the first step and LM in the second step. ICB stands for individual-cluster-based robust inference. RoM is linear mixed model with family-cluster-based robust inference.

**Table 1:**
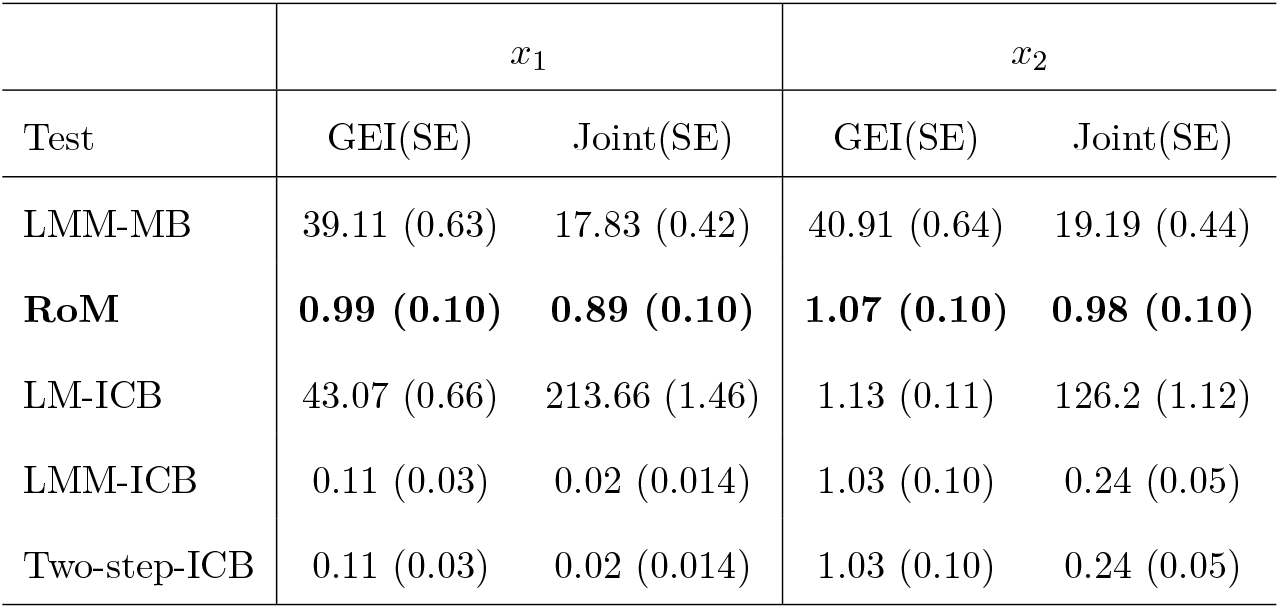
Empirical type I error rates for heritable environmental exposure *x*_1_ and non-heritable environmental exposure *x*_2_ divided by genome-wide significant level 5 × 10^−8^. LMM stands for linear mixed model. LM stands for linear model. ICB stands for individual-cluster-based robust inference. RoM is linear mixed model with family-cluster-based robust inference.

### 3.2 Application to family data and longitudinal data

Figure 2 and Figure 3 present the QQ plots of genetic marginal test, GEI test and joint test of RoM and five alternative models, applied to family data from FHS and longitudinal data from ARIC, respectively. The alternative models explored various aspects, including the functional form of BMI exposure, the inclusion of a random effect for BMI, and the choice between model-based or robust association tests.

**Figure 2:**
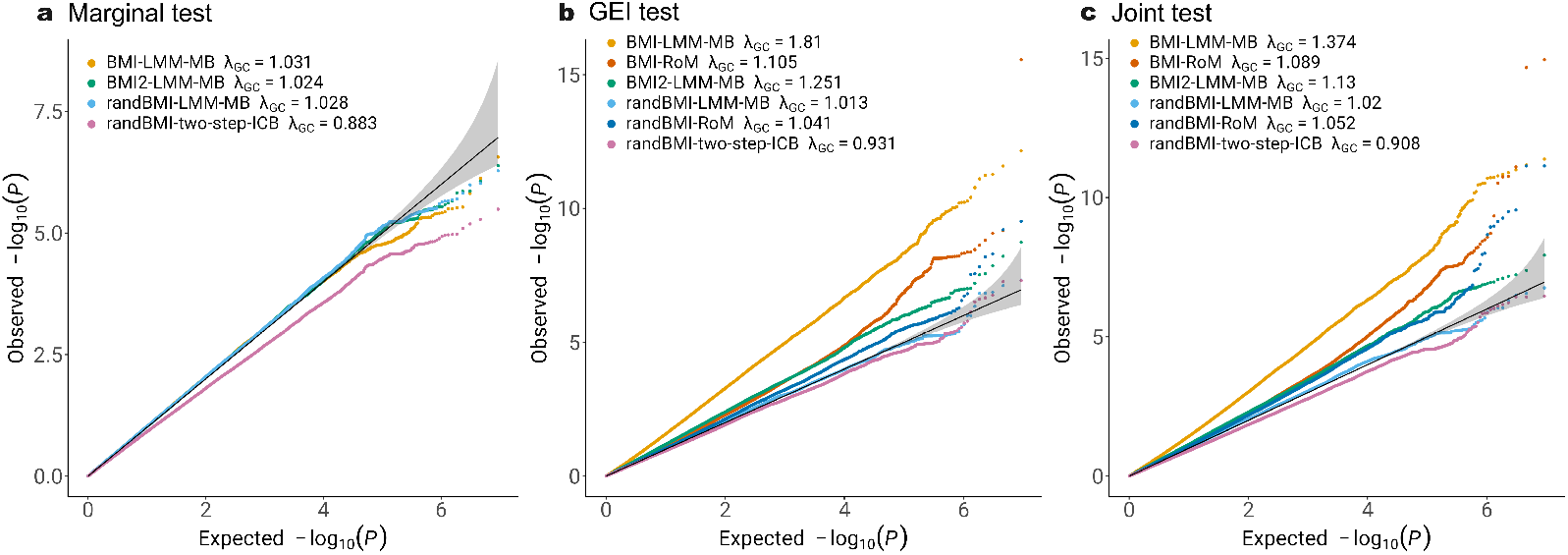
Comparison of RoM and five alternative models in FHS study of gene-BMI interaction on WHR. RoM marginal test result is not shown, as it is identical to that from LMM-MB. BMI2 stands for the null model including quadratic BMI terms. randBMI stands for the model including random slope of BMI. LMM stands for linear mixed model. LM stands for linear model. Two-step stands for two-step approach applying LMM in the first step and LM in the second step. ICB stands for individual-cluster-based robust inference. RoM is linear mixed model with family-cluster-based robust inference.

**Figure 3:**
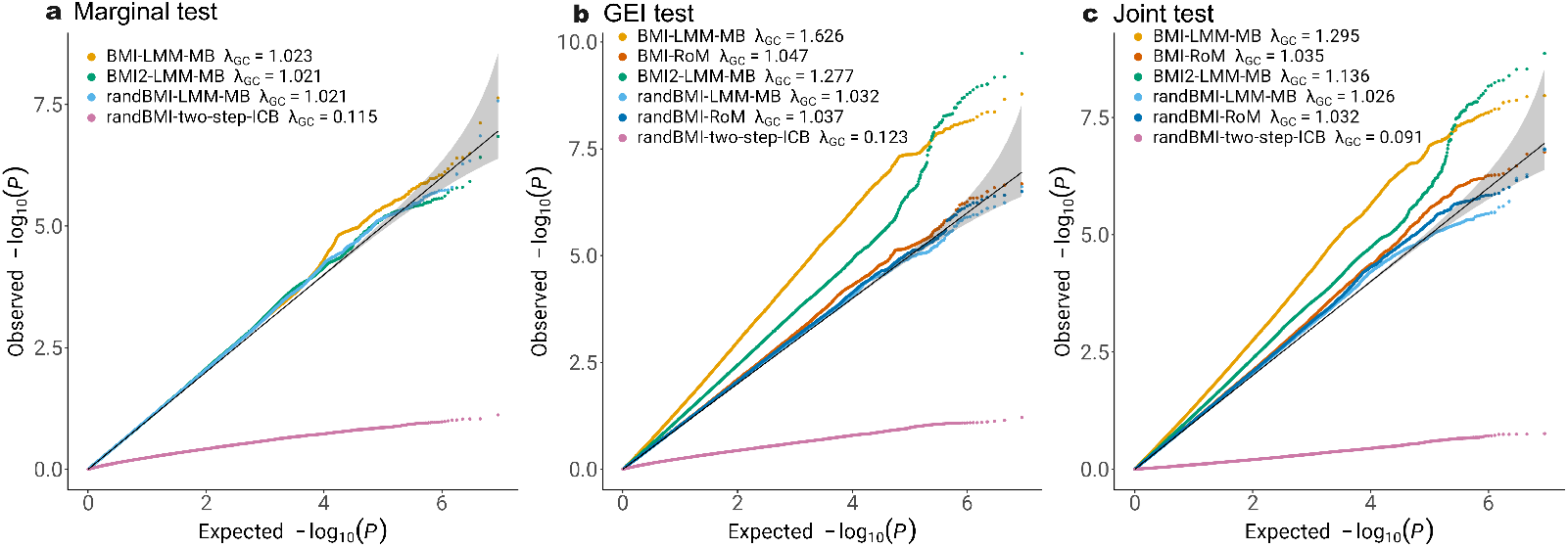
Comparison of RoM and five alternative models in ARIC study of gene-BMI interaction on WHR. RoM marginal test result is not shown, as it is identical to that from LMM-MB. BMI2 stands for the null model including quadratic BMI terms. randBMI stands for the model including random slope of BMI. LMM stands for linear mixed model. LM stands for linear model. Two-step stands for two-step approach applying LMM in the first step and LM in the second step. ICB stands for individual-cluster-based robust inference. RoM is linear mixed model with family-cluster-based robust inference.

Figure 2 shows that there are factors other than the nonlinear BMI effect that contribute to the inflation of type I error in FHS with family data. Using random slope for BMI appears to be capable of addressing the problem of inflated type I error. RoM applied to the mixed effect model with random slope of BMI also gives reasonable p values. It is worth noting that even in the model where random slope of BMI is considered, type I error rates with two-step approach is deflated in marginal test, GEI test, and Joint test (Marginal *λ*_*GC*_ = 0.883; GEI *λ*_*GC*_ = 0.931; Joint *λ*_*GC*_ = 0.908) which is consistent with the findings in simulation study.

Figure 3 shows that model misspecification also leads to inflated GEI tests in ARIC study with longitudinal data, and RoM is able to fix type I error inflation when treating repeated measurements of the same individual as a cluster. The type I error rates is dramatically deflated with the two-step approach when treating each measurement independently in the second step.

### 3.3 Application to Biobank-scale data and signal identification

We compared association results with and without the inclusion of related participants (Figure 4 and Figure 5). Overall, we observed that datasets with larger sample sizes tended to yield more significant associations (Panel c of Figure 4 and Figure 5). We identified 51 loci from interaction test (Supplementary Table S1) and 240 loci from joint test (Supplementary Table S2) at genome-wide significance level of 5 × 10^−8^ from samples with inclusion of related participants. For independent participants, we identified 34 loci from interaction test (Supplementary Table S3) and 143 loci from joint test (Supplementary Table S4). Our findings reveal that most loci are more significant than those reported by Westerman et al. (2021). In interaction test, RoM identified the locus 2:165349902-165846592 on chromosome 2 with rs1128249 (*P* = 9.86 × 10^−81^) as its index SNP. Nearby genes are *GRB14, COBLL1, SLC38A11*. The genes mentioned were all identified in previous work in unrelated samples (Westerman et al. 2021) (Supplementary Table S1). We also annotated lead SNPs from joint test, and identified the top locus 6:126278230-127529780 on chromosome 6 with rs72961013 (*P* = 1.14 × 10^−186^) as index SNP. Nearby genes are *TRMT11, CENPW*, and *RSPO3*. Gene *VEGFA* was also identified by RoM on locus 6:43753325-43828582 with rs4711750 (*P* = 4.79 × 10^−131^) as index SNP (Supplementary Table S2). Supplementary Figure S2 and Figure S3 generated by LocusZoom web tool (Boughton et al. 2021) shows the hits in GWAS catalog traits waist-to-hip ratio.

**Figure 4:**
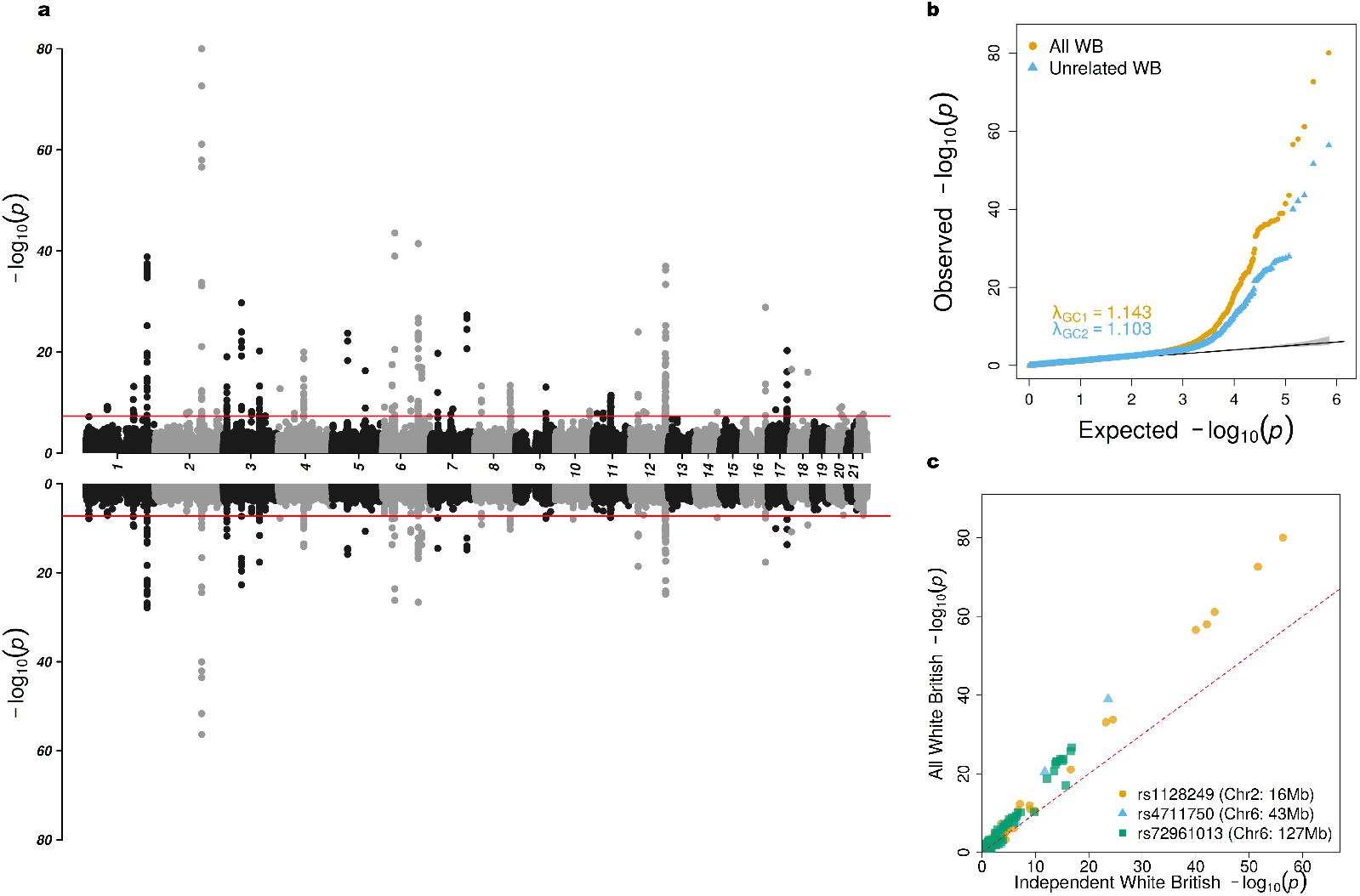
Comparison of White British participants in UK Biobank gene-sex interaction test of WHR. a) Miami plot showing results for all White British participants (top) and independent White British participants (bottom). b) QQ plot of all White British in orange and independent White British in blue. c) Scatter plot of p-values on the -log10 scale between all White British versus independent White British colored by three selected loci.

**Figure 5:**
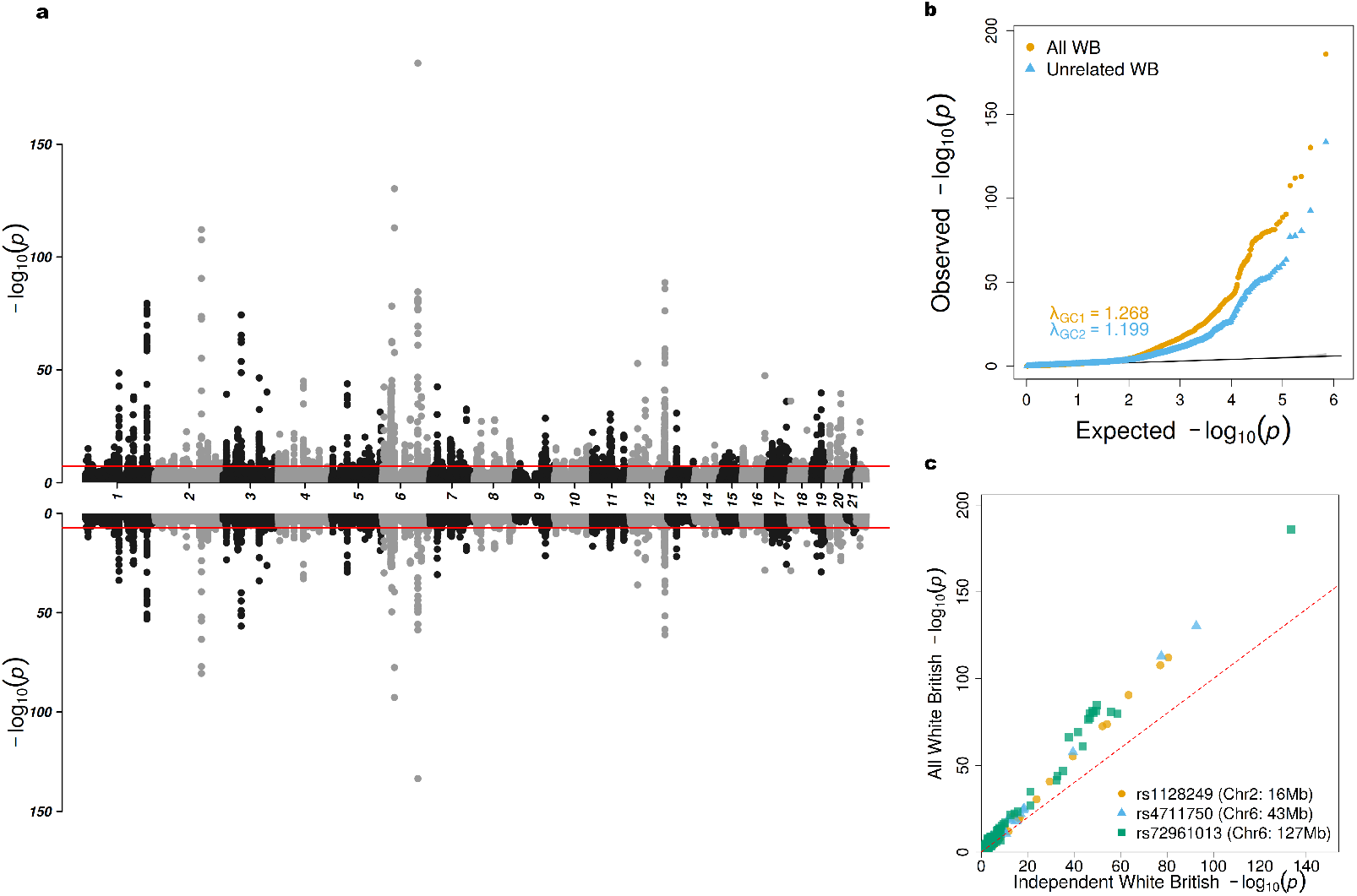
Comparison of White British participants in UK Biobank gene-sex joint test of WHR. a) Miami plot showing results for all White British participants (top) and independent White British participants (bottom). b) QQ plot of all White British in orange and independent White British in blue. c) Scatter plot of p-values on the -log10 scale between all White British versus independent White British colored by three selected loci.

## 4 Discussion

In this study, we addressed two prevalent challenges in large-scale Genome-Wide Association Studies (GWAS) involving related samples: model misspecification and computational efficiency. These issues are particularly salient in Gene-Environment Interaction (GEI) studies, where missing non-linear environmental exposures can introduce biases. Through both simulations and real data applications, we demonstrated that the bias lead to inflated type I error. To address this issue, we developed RoM, a robust inference method that employs a sandwich estimator within the framework of linear mixed-effects models. The sandwich estimator is advantageous as it ensures that variance estimates remain asymptotically normal under regular conditions and provides consistent standard error estimates even in the presence of model misspecification, which was proved by our simulation results (Figure 1). The asymptotic properties of the sandwich estimator imply that as the sample size increases, the estimates converge to the true variance, thereby enhancing reliability. In our simulation study with a sample size of 8,500 (Supplementary Figure S4), we observed inflated tails in the QQ plots; however, with an increased sample size of 85,000, the tails were closer to 45-degree line. RoM also shows efficiency in handling large-scale genomic data, with the time complexity of robust variance computation being *O*(*N*). RoM completed a scan of 1 million SNPs in approximately 1.52 CPU hours for a sample size of 8,500 and 9.8 CPU hours for a sample size of 85,000 using a single-threaded configuration on Intel(R) Xeon(R) Gold 6254 CPU @ 3.10GHz.

Further simulation analysis revealed that the commonly employed two-step approach can lead to deflated p-values in GEI and joint tests for heritable environmental exposures. This loss of power is primarily due to misspecified variance estimates in the linear model used in the second step. We also observed the same results in strategy LMM-ICB, and the deflation is caused by the misspecified clustering. Importantly, the deflation occurs when the environmental exposure is heritable. In this case, researchers should be cautious about model misspecification when using the two-step approach for heritable exposures, since the inflation due to misspecification, combined with the deflation resulting from the two-step approach, may yield QQ plots that appear normal.

We applied RoM to FHS and ARIC study data to explore the association between gene and WHR with gene-BMI interaction. Our findings indicated that RoM effectively accounts for the correlation structures typically encountered in repeated measures and provides well-calibrated p values in the context of longitudinal data. However, we observed that RoM did not sufficiently control type I error inflation in FHS family data. Two potential factors may contribute to it. First, there may be additional variables beyond the nonlinear effects of BMI that cause type I error inflation. The FHS results suggest the incorporation of a random slope for BMI appears to address this issue. Second, the relatively small sample sizes in FHS employed in this analysis may further limit the robustness of sandwich estimators.

In the gene-sex interaction study of WHR using UK Biobank data, we applied the RoM approach to evaluate its efficacy in identifying significant associations within a large cohort. The results highlighted that larger sample sizes generally yielded more significant associations, aligning with the theory that greater statistical power can be achieved by increasing sample size. Importantly, it emphasizes the potential power increasing benefits of including related individuals in genetic studies. Additionally, there is a positive correlation between sample size and GIF (Westerman et al. 2021). Despite a 50% increase in sample size for the all White British sample relative to the independent White British sample, RoM successfully produced well-calibrated p values. This is evidenced by the comparable genomic inflation factors observed for the interaction test (1.143 versus 1.103) and the joint test (1.268 versus 1.199).

In summary, RoM is a computationally efficient tool designed for conducting single-variant GWAS that effectively handles gene-environment interactions in datasets comprising millions of related individuals or observations. RoM allows for the incorporation of multiple environmental exposures and gene by environment interactions while providing robust standard error estimates for clustered data.

## Data Availability

All data produced in the present study are available upon reasonable request to the authors.

https://github.com/MengyuZhang1307/RoM

## 5 Acknowledgments

This research was conducted using the UK Biobank Resource under Application Numbers 42646 and 92681. The Atherosclerosis Risk in Communities study has been funded in whole or in part with Federal funds from the National Heart, Lung, and Blood Institute, National Institutes of Health, Department of Health and Human Services, under Contract nos. (75N92022D00001, 75N92022D00002, 75N92022D00003, 75N92022D00004, 75N92022D00005). The authors thank the staff and participants of the ARIC study for their important contributions. Funding was also supported by R01HL087641 and R01HL086694; National Human Genome Research Institute contract U01HG004402; and National Institutes of Health contract HHSN268200625226C. Infrastructure was partly supported by Grant Number UL1RR025005, a component of the National Institutes of Health and NIH Roadmap for Medical Research. This research was conducted in part using data and resources from the Framingham Heart Study of the National Heart Lung and Blood Institute of the National Institutes of Health and Boston University School of Medicine. The Framingham Heart Study (FHS) acknowledges the support of contracts NO1-HC-25195, HHSN268201500001I, and 75N92019D00031 from the National Heart, Lung, and Blood Institute for this research. We also acknowledge the dedication of the FHS study participants without whom this research would not be possible. The authors acknowledge the Texas Advanced Computing Center (TACC) at The University of Texas at Austin for providing computational resources that have contributed to the simulation results reported within this paper. URL: http://www.tacc.utexas.edu.

This work was supported by National Institutes of Health grant R01HL145025, R01DK122503, and R01HL156991.

## SUPPLEMENTARY MATERIAL

### R-package RoM

RoM is an R package for Mixed-model association test (available at https://github.com/MengyuZhang1307/RoM). Perform generalized linear mixed robust single-variant gene-environment interaction tests and joint tests for common variants.

### Supplementary Tables

Supplementary Tables contains the results of significant loci identified from the interaction/joint test in the UK Biobank analysis of Waist-Hip Ratio from white British samples.

### Supplementary Notes

Supplementary Notes contains the full derivation of RoM and supplementary figures.

