## Supplementary Notes and Supplementary Figures for "Robust Mixed Model Association Test for Gene-Environment Interactions"

### A Derivation of Robust Test

Considering full mixed model Equation (1)

$$E(Y_i) = \boldsymbol{X}_i\boldsymbol{\alpha} + g_i\beta + \boldsymbol{K}_i\boldsymbol{\gamma} + r_i, \tag{1}$$

the scaled residual is

$$\begin{aligned}\boldsymbol{R} &= \hat{\boldsymbol{V}}^{-1}(\boldsymbol{Y} - \boldsymbol{X}\hat{\boldsymbol{\alpha}} - \boldsymbol{g}\hat{\beta} - \boldsymbol{K}\hat{\boldsymbol{\gamma}} - \hat{\boldsymbol{r}}) \\ &= \hat{\boldsymbol{\Sigma}}^{-1}(\boldsymbol{Y} - \boldsymbol{X}\hat{\boldsymbol{\alpha}} - \boldsymbol{g}\hat{\beta} - \boldsymbol{K}\hat{\gamma})\end{aligned} \tag{2}$$

where $\hat{\boldsymbol{V}} = \hat{\phi}\boldsymbol{I_N}$ and $\hat{\phi}$ is estimated dispersion parameter. $\hat{\boldsymbol{\Sigma}} = \hat{\boldsymbol{V}} + \sum_{l=1}^{L}\hat{\lambda}_l\boldsymbol{\Psi}_l$ is estimated covariance matrix for samples from full model, where $\hat{\lambda}_l$ is estimated variance component for $L$ random effect and $\boldsymbol{\Psi}_l$ is a $N \times N$ kinship matrix or relationship matrix. Projection matrix $\hat{\boldsymbol{P}} = \hat{\boldsymbol{\Sigma}}^{-1} - \hat{\boldsymbol{\Sigma}}^{-1}\boldsymbol{X}(\boldsymbol{X}^T\hat{\boldsymbol{\Sigma}}^{-1}\boldsymbol{X})^{-1}\boldsymbol{X}^T\hat{\boldsymbol{\Sigma}}^{-1}$.

Now considering null model in Equation (3)

$$E(Y_i) = \boldsymbol{X}_i\boldsymbol{\alpha} + r_i, \tag{3}$$

scaled residuals are computed as

$$\begin{aligned}\tilde{\boldsymbol{R}} &= \hat{\hat{\boldsymbol{V}}}^{-1}(\boldsymbol{Y} - \boldsymbol{X}\hat{\hat{\boldsymbol{\alpha}}} - \hat{\hat{\boldsymbol{r}}}) \\ &= \hat{\hat{\boldsymbol{\Sigma}}}^{-1}(\boldsymbol{Y} - \boldsymbol{X}\hat{\hat{\boldsymbol{\alpha}}}) \\ &= \hat{\hat{\boldsymbol{\Sigma}}}^{-1}(\boldsymbol{Y} - \boldsymbol{X}(\boldsymbol{X}^T\hat{\hat{\boldsymbol{\Sigma}}}^{-1}\boldsymbol{X})^{-1}\boldsymbol{X}^T\hat{\hat{\boldsymbol{\Sigma}}}^{-1}\boldsymbol{Y})\end{aligned} \tag{4}$$

$\hat{\hat{\boldsymbol{P}}} = \hat{\hat{\boldsymbol{\Sigma}}}^{-1} - \hat{\hat{\boldsymbol{\Sigma}}}^{-1}\boldsymbol{X}(\boldsymbol{X}^T\hat{\hat{\boldsymbol{\Sigma}}}^{-1}\boldsymbol{X})^{-1}\boldsymbol{X}^T\hat{\hat{\boldsymbol{\Sigma}}}^{-1}$ where $\hat{\hat{\boldsymbol{V}}} = \hat{\hat{\phi}}\boldsymbol{I_N}$, and $\hat{\hat{\boldsymbol{\Sigma}}}^{-1}$ is inverse of the estimated covariance matrix for samples from null model. With the assumption that true values of $\beta$ and $\boldsymbol{\gamma}$ are small, the variance component estimates for $\phi$ and $\boldsymbol{\Psi}$ from Equation (3) are not dramatically different from the estimates from Equation (1), so $\hat{\hat{\boldsymbol{\Sigma}}} \approx \hat{\boldsymbol{\Sigma}}$ and $\hat{\hat{\boldsymbol{\Sigma}}}^{-1} - \hat{\hat{\boldsymbol{\Sigma}}}^{-1}\boldsymbol{X}(\boldsymbol{X}^T\hat{\hat{\boldsymbol{\Sigma}}}^{-1}\boldsymbol{X})^{-1}\boldsymbol{X}^T\hat{\hat{\boldsymbol{\Sigma}}}^{-1} = \hat{\hat{\boldsymbol{P}}} \approx \hat{\boldsymbol{P}}$. Therefore, $\tilde{\boldsymbol{R}} \approx \hat{\boldsymbol{P}}\boldsymbol{Y}$.

The "bread" matrix for joint test can be approximated by

$$\begin{aligned}\hat{\boldsymbol{B}}_J &= (\boldsymbol{W}^T\hat{\hat{\boldsymbol{P}}}\boldsymbol{W})^{-1} \\ &= \left(\boldsymbol{W}^T\hat{\hat{\boldsymbol{\Sigma}}}^{-1}\boldsymbol{W} - \boldsymbol{W}^T\hat{\hat{\boldsymbol{\Sigma}}}^{-1}\boldsymbol{X}(\boldsymbol{X}^T\hat{\hat{\boldsymbol{\Sigma}}}^{-1}\boldsymbol{X})^{-1}\boldsymbol{X}^T\hat{\hat{\boldsymbol{\Sigma}}}^{-1}\boldsymbol{W}\right)^{-1} \\ &= \begin{bmatrix}\boldsymbol{g}^T\hat{\hat{\boldsymbol{P}}}\boldsymbol{g} & \boldsymbol{g}^T\hat{\hat{\boldsymbol{P}}}\boldsymbol{K} \\ \boldsymbol{K}^T\hat{\hat{\boldsymbol{P}}}\boldsymbol{g} & \boldsymbol{K}^T\hat{\hat{\boldsymbol{P}}}\boldsymbol{K}\end{bmatrix}^{-1}\end{aligned} \tag{5}$$

$\hat{\boldsymbol{B}}_J$ is inverse of Schur complement of the block $\boldsymbol{X}^T\hat{\hat{\boldsymbol{\Sigma}}}^{-1}\boldsymbol{X}$ of the matrix $\boldsymbol{B}$

$$\boldsymbol{B} = \begin{bmatrix}\boldsymbol{X}^T\hat{\hat{\boldsymbol{\Sigma}}}^{-1}\boldsymbol{X} & \boldsymbol{X}^T\hat{\hat{\boldsymbol{\Sigma}}}^{-1}\boldsymbol{W} \\ \boldsymbol{W}^T\hat{\hat{\boldsymbol{\Sigma}}}^{-1}\boldsymbol{X} & \boldsymbol{W}^T\hat{\hat{\boldsymbol{\Sigma}}}^{-1}\boldsymbol{W}\end{bmatrix}^{-1} \tag{6}$$

Effect sizes of genetic effect and interaction effect is approximated by

$$\hat{\boldsymbol{\zeta}} = \hat{\boldsymbol{B}}_J \boldsymbol{W}^T \tilde{\boldsymbol{R}} = (\boldsymbol{W}^T \hat{\hat{\boldsymbol{P}}} \boldsymbol{W})^{-1} \boldsymbol{W}^T \tilde{\boldsymbol{R}} \tag{7}$$

The "bread" matrix for interaction test $\boldsymbol{B}_I$

$$\hat{\boldsymbol{B}}_I = (\boldsymbol{K}^T \hat{\hat{\boldsymbol{H}}} \boldsymbol{K})^{-1} = \left( \boldsymbol{K}^T \hat{\hat{\boldsymbol{P}}} \boldsymbol{K} - \boldsymbol{K}^T \hat{\hat{\boldsymbol{P}}} \boldsymbol{g} (\boldsymbol{g}^T \hat{\hat{\boldsymbol{P}}} \boldsymbol{g})^{-1} \boldsymbol{g}^T \hat{\hat{\boldsymbol{P}}} \boldsymbol{K} \right)^{-1} \tag{8}$$

where $\hat{\hat{\boldsymbol{H}}} = \hat{\hat{\boldsymbol{P}}} - \hat{\hat{\boldsymbol{P}}} \boldsymbol{g} (\boldsymbol{g}^T \hat{\hat{\boldsymbol{P}}} \boldsymbol{g})^{-1} \boldsymbol{g}^T \hat{\hat{\boldsymbol{P}}}$ is a $N \times N$ projection matrix. Model-based GEI test statistic is

$$t_I = \hat{\boldsymbol{\gamma}}^T \hat{\boldsymbol{B}}_I^{-1} \hat{\boldsymbol{\gamma}} \tag{9}$$

Model-based joint test statistic is

$$t_J = \hat{\boldsymbol{\zeta}}^T \hat{\boldsymbol{B}}_J^{-1} \hat{\boldsymbol{\zeta}} \tag{10}$$

Residual of $\boldsymbol{W}$ is $\tilde{\boldsymbol{W}} \approx \boldsymbol{W} - \boldsymbol{X}(\boldsymbol{X}^T \hat{\hat{\boldsymbol{\Sigma}}}^{-1} \boldsymbol{X})^{-1} \boldsymbol{X}^T \hat{\hat{\boldsymbol{\Sigma}}}^{-1} \boldsymbol{W}$. Scaled residuals after adjusting for genotypes and interaction term are computed by

$$\begin{aligned} \tilde{\tilde{\boldsymbol{R}}} &= \tilde{\boldsymbol{R}} - \hat{\hat{\boldsymbol{P}}} \boldsymbol{W} (\boldsymbol{W}^T \hat{\hat{\boldsymbol{P}}} \boldsymbol{W})^{-1} \boldsymbol{W}^T \tilde{\boldsymbol{R}} \\ &\approx \left( \hat{\boldsymbol{P}} - \hat{\boldsymbol{P}} \boldsymbol{W} (\boldsymbol{W}^T \hat{\boldsymbol{P}} \boldsymbol{W})^{-1} \boldsymbol{W}^T \hat{\boldsymbol{P}} \right) \boldsymbol{Y} \end{aligned} \tag{11}$$

A $1 \times (q+1)$ score vector for subject $i$ can be constructed by

$$\tilde{\boldsymbol{S}}_{J,i} = \tilde{\boldsymbol{W}}_i \tilde{\tilde{\boldsymbol{R}}}_i \tag{12}$$

Joint test "meat" matrix adjusted for clustering is Equation (13).

$$\hat{\boldsymbol{M}}_J = \sum_{j=1}^{N_c} \left[ \sum_{i \in c_j} \tilde{\boldsymbol{S}}_{J,i} \right]^T \left[ \sum_{i \in c_j} \tilde{\boldsymbol{S}}_{J,i} \right] \tag{13}$$

The "meat" matrix for interaction test $\hat{\boldsymbol{M}}_I$ is the corresponding block of $\hat{\boldsymbol{M}}_J$.

### B Computational efficiency

Hypothesis test for common variants is conducted independently for each variant and , so they can be parallelized in batches, and multiple variants can be tested at the same time.

Considering a situation where batch size is $n$ and with $q$ environment factors of interests. Then we have a new $N \times (n + nq)$ design matrix for a certain batch $b$.

$$\boldsymbol{W}^{(b)} = (\boldsymbol{g}_1, ..., \boldsymbol{g}_n, \boldsymbol{g}_1 \boldsymbol{E}_1, \boldsymbol{g}_2 \boldsymbol{E}_1, ..., \boldsymbol{g}_n \boldsymbol{E}_1, ..., \boldsymbol{g}_1 \boldsymbol{E}_q, \boldsymbol{g}_2 \boldsymbol{E}_q, ..., \boldsymbol{g}_n \boldsymbol{E}_q)$$

$\tilde{\tilde{\boldsymbol{R}}}$ in Equation (11) is then expanded to a $N \times n$ matrix $\tilde{\tilde{\boldsymbol{R}}}^{(b)}$. Each column is the residuals for each variant accordingly. It is worthy to mention that the computational complexity of "meat" matrix is $O(N)$ given batch sizes.

$$\tilde{\tilde{\boldsymbol{R}}}^{(b)} = \tilde{\boldsymbol{R}} - \hat{\hat{\boldsymbol{P}}} \boldsymbol{W}^{(b)} \left( \left( \boldsymbol{j}_{1+q} \otimes \boldsymbol{I}_n \right) \odot \left( \left( \boldsymbol{W}^{(b)T} \hat{\hat{\boldsymbol{P}}} \boldsymbol{W}^{(b)} \odot (\boldsymbol{J}_{1+q} \otimes \boldsymbol{I}_n) \right)^{-1} \boldsymbol{W}^{(b)T} \tilde{\boldsymbol{R}} \right) \right)$$

Residual $\boldsymbol{W}$ becomes a $N \times (n + nq)$ matrix $\tilde{\boldsymbol{W}}^{(b)} \approx \boldsymbol{W}^{(b)} - \boldsymbol{X}(\boldsymbol{X}^T \hat{\tilde{\boldsymbol{\Sigma}}}^{-1} \boldsymbol{X})^{-1} \boldsymbol{X}^T \hat{\tilde{\boldsymbol{\Sigma}}}^{-1} \boldsymbol{W}^{(b)}$. The score vector for joint test is

$$\tilde{\boldsymbol{S}}_J^{(b)} = \tilde{\boldsymbol{W}}^{(b)} \odot \tilde{\tilde{\boldsymbol{R}}}^{(b)} \tag{14}$$

The "meat" matrix for SNP batch $b$ is

$$\hat{\boldsymbol{M}}_J^{(b)} = \tilde{\boldsymbol{S}}_J^{(b)} \boldsymbol{J}_{sparse} \boldsymbol{J}_{sparse}^T \left( \tilde{\boldsymbol{S}}_J^{(b)} \right)^T \tag{15}$$

Membership matrix $\boldsymbol{J}_{sparse}^T$ is a $N \times N_c$ sparse matrix. In this matrix, the entry at the $i$-th row and $j$-th column is 1 if individual $i$ belongs to cluster $j$; otherwise, the entry is 0.

### C Supplementary Figures

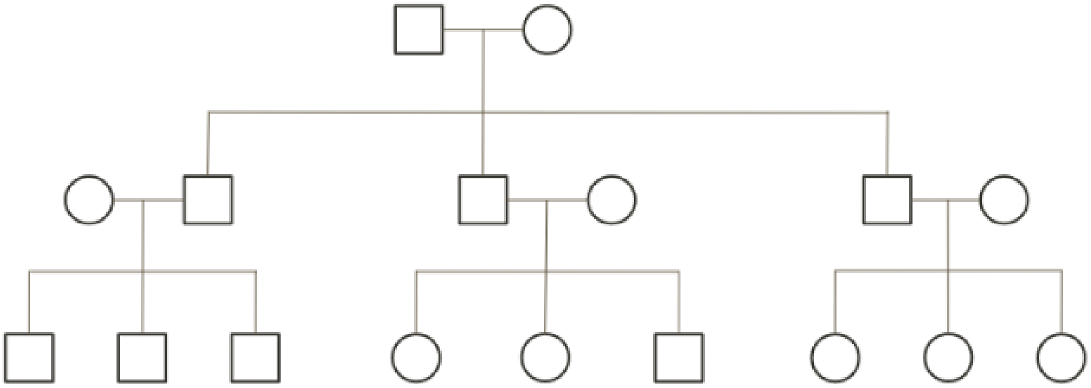

Figure S1: Pedigree of extended families.

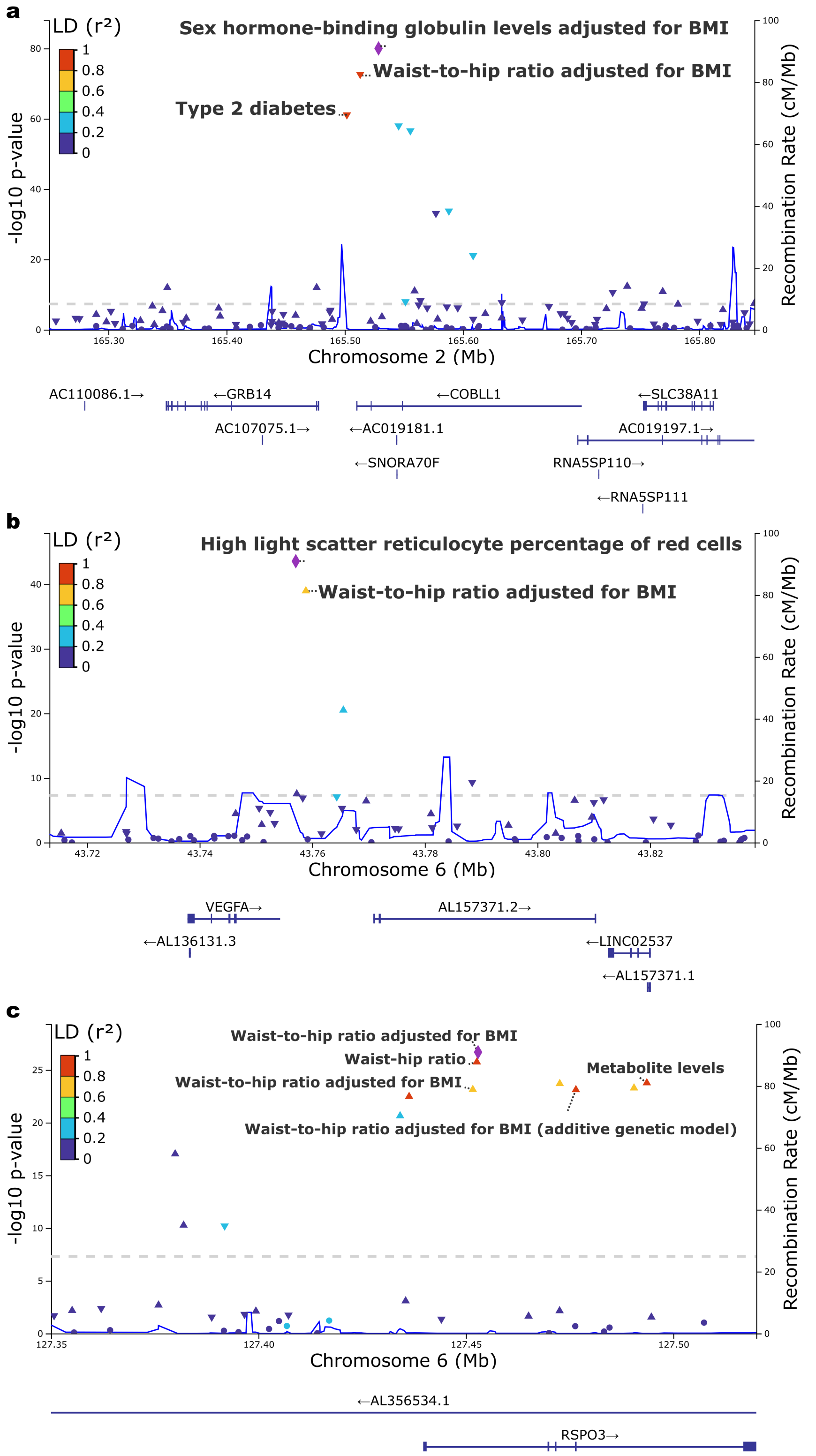


Figure S2: Regional plot of significant loci for gene-sex interaction test of WHR in UK Biobank of all White British. a) Locus on Chromosome 2 labeled with catalog traits. b-c) Locus on Chromosome 6 labeled with catalog traits.

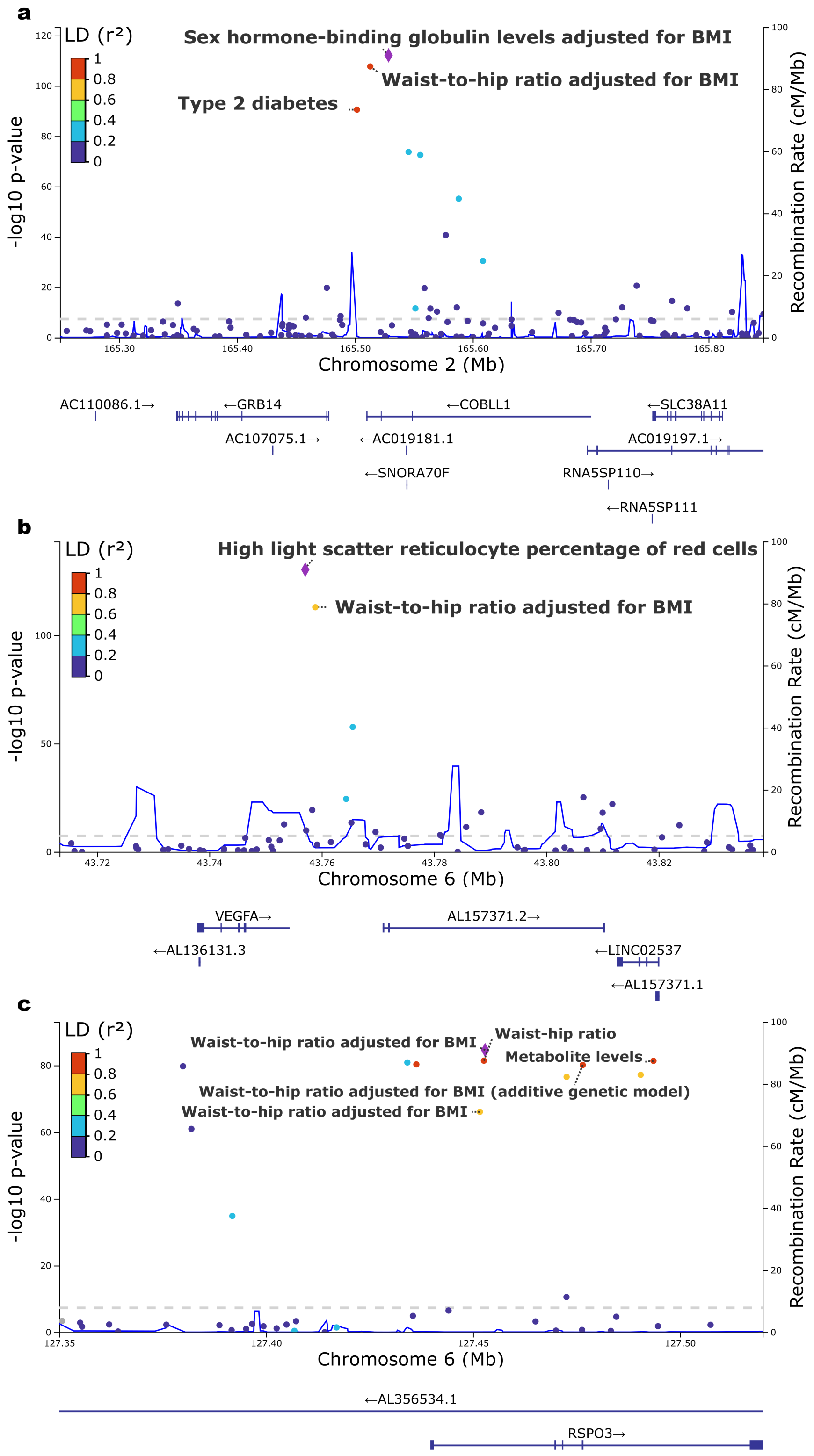


Figure S3: Regional plot of significant loci for gene-sex joint test of WHR in the UK Biobank of all White British. Variants with -log10(P) > 120 were removed. a) Locus on Chromosome 2 labeled with catalog traits. b-c) Locus on Chromosome 6 labeled with catalog traits.

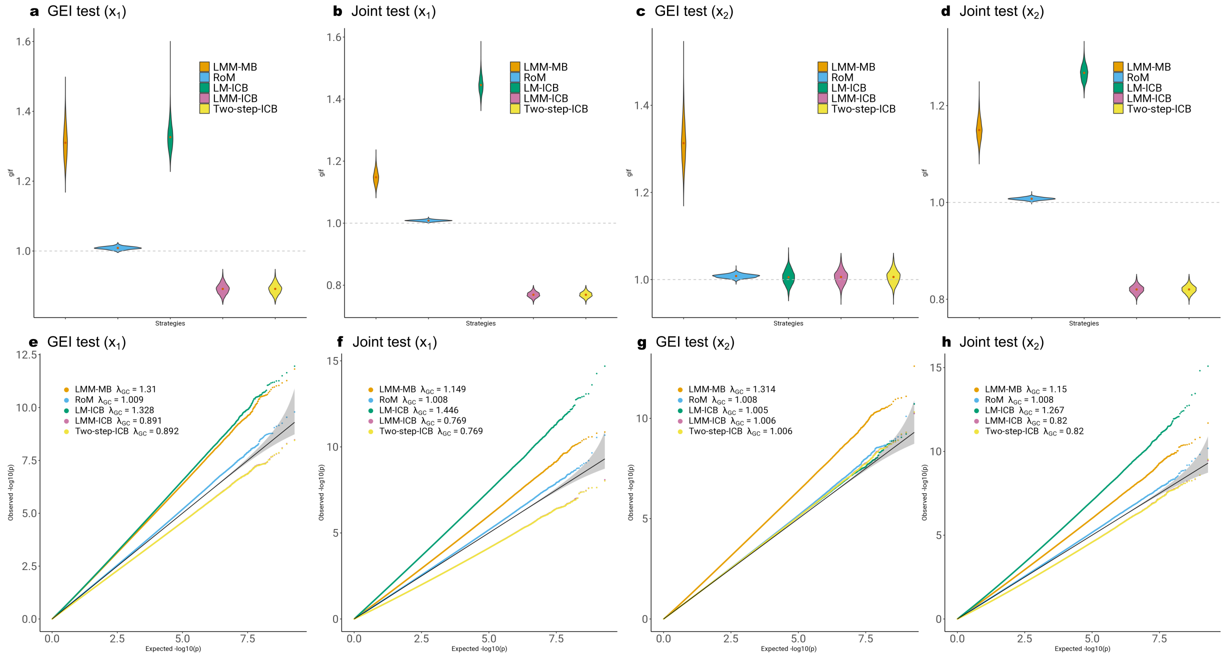


Figure S4: Comparison of GIF and QQ plots of RoM and four alternative strategies under the null hypothesis of no genetic effect or GEI effect for a sample size of 8,500. a-d) Genomic inflation factors over 2,000 replicates. e-h) QQ plots of 2 billion SNPS from 2,000 replicates combined. LMM stands for linear mixed model. LM stands for linear model. Two-step stands for two-step approach applying LMM in the first step and LM in the second step. ICB stands for individual-cluster-based robust inference. RoM is linear mixed model with family-cluster-based robust reference.
